# Modeling the use of SARS-CoV-2 vaccination to safely relax non-pharmaceutical interventions

**DOI:** 10.1101/2021.03.12.21253481

**Authors:** Alicia N.M. Kraay, Molly E. Gallagher, Yang Ge, Peichun Han, Julia M. Baker, Katia Koelle, Andreas Handel, Benjamin A Lopman

## Abstract

In response to the COVID-19 pandemic, widespread non-pharmaceutical interventions (NPIs), including physical distancing, mask wearing, and enhanced hygiene, have been implemented. As of March 2021, three effective vaccines have been approved for emergency use in the United States, with several other vaccines in the pipeline. We use a transmission model to study when and how NPIs could be relaxed in the United States with relative safety as vaccination becomes more widespread. We compare different relaxation scenarios where NPIs begin to relax 0-9 months after vaccination begins for both a one dose and two dose strategy, with historical levels of social interactions being reached within 1 month to 1 year. In our model, vaccination can allow widespread relaxation of NPIs to begin safely within 2 to 9 months, greatly reducing deaths and peak health system burden compared to relaxing NPIs without vaccination. Vaccinated individuals can safely begin to relax NPIs sooner than unvaccinated individuals. The extent of delay needed to safely reopen depends primarily on the rate of vaccine rollout, with the degree of protection against asymptomatic infection playing a secondary role. If a vaccination rate of 3 million doses/day can be achieved, similar to the typical rollout speed of seasonal influenza vaccination, NPIs could begin to be safely relaxed in 2-3 months. With a vaccination rate of 1 million doses/day, a 6–9-month delay is needed. A one dose strategy is preferred if relative efficacy is similar to a two-dose series, but the relative benefit of this strategy is minimal when vaccine rollout is fast. Due to the urgent need to pursue strategies that enable safe relaxation of NPIs, we recommend a two-dose strategy with an initial delay of at least 3 months in relaxing restrictions further, and that the speed of vaccine rollout be given immediate priority.

## Introduction

The COVID-19 pandemic has caused catastrophic loss of life and health system strain in the United States, with 506,373 deaths as of March 3, 2021 [1]. Widespread non-pharmaceutical interventions (NPIs) initially reduced the spread [2]. However, such interventions have significant social and economic costs and are not sustainable in the long-term. As a result, these NPIs have been slowly relaxed which has led to increased community transmission [3]. In fall and winter 2020, cases began to rise again, leading to renewed restrictions in many parts of the country in an effort to slow transmission.

At the same time, vaccine development has been proceeding at a rapid pace, with three vaccines approved for emergency use in the United States as of March 2021, about one year after the first cases occurred in the United States [4, 5, 6]. Two mRNA vaccines, one by Pfizer/BioNTech and one by Moderna, are given in a 2-dose regimen and have efficacy against symptomatic SARS-CoV-2 of 90% or greater. The most recently approved Johnson and Johnson vaccine is given as a single dose and has a somewhat lower efficacy (66% against symptomatic SARS-CoV-2 and 85% against severe disease) [7, 8, 6]. As vaccination is rolled out in the United States, there is an imperative to relax social distancing and other NPIs without causing a resurgence in transmission.

Due to the initially limited supply, using one dose instead of two for the mRNA vaccines has been discussed as a potential strategy to allow more people to be vaccinated more quickly [9, 10]. Such a strategy has previously been used during a yellow fever epidemic [11] and has been considered as a possibility for pandemic flu [12]. In general, the utility of this strategy depends on both the baseline transmission rate, the relative performance of 1 vs. 2 doses, and the vaccine mechanism of action [12], which is presently unknown for SARS-CoV-2. The UK has already chosen this strategy, and top officials in the United States are discussing this possibility as well as other modified dosing schedules. We therefore explicitly model the potential use of a one dose strategy to extend supply in a context where non-pharmaceutical interventions are also being relaxed.

The potential impact of SARS-CoV-2 vaccination depends not only on individual protection against severe disease and/or mortality, but also the indirect effects of vaccination, which act on transmission. Higher levels of indirect effects could greatly enhance the population level impact of vaccination [13, 14]. In clinical trials for SARS-CoV-2, efficacy has been measured against symptomatic COVID-19, which is determined by both protection against infection and subsequent protection against severe disease if infected [15]. Less data is available on protection against infection and transmission, both of which influence the strength of indirect effects and benefits for unvaccinated individuals. Given both the very high efficacy of currently used SARS-CoV-2 vaccines and promising initial data from field studies, some protection against infection is likely, but its degree is unknown [16, 17].

Given that vaccinated individuals are likely to have a reduced likelihood of infection, NPIs might be able to be safely relaxed sooner for this group, particularly for younger adults who are not at high risk of severe disease. In practice, individuals are likely to begin engaging in higher risk behavior soon after completing their vaccine series. Indeed, the Centers for Disease Control and Prevention (CDC) is already recommending the certain activities are safe for fully vaccinated people [18]. We therefore explicitly model preferential relaxation of NPIs for vaccinated compared with unvaccinated individuals, exploring how this change influences population risk.

In this analysis, we study whether and how non-pharmaceutical interventions can be relaxed safely. We assess whether vaccinated individuals might be able to regain their social contacts more quickly without greatly increasing population transmission risk. To explore potential trade-offs in speed and efficacy, we compare a two dose vaccination strategy for the two mRNA vaccines with the potential impacts if a 1-dose strategy were used. We do not consider the Johnson and Johnson vaccine separately, as most of the initial vaccine supply is focused on the mRNA vaccines, but the potential impact of the Johnson and Johnson vaccine is approximated by our one dose scenarios with high efficacy. We also explore how the vaccine mechanism of action, including the extent to which vaccination reduces infection (and therefore, the level of indirect effects), affects overall impacts.

## Methods

### Model structure

Our base model includes seven compartments. Initially, most individuals are susceptible to infection (*S*). Upon exposure, they enter a latent period (*E*), during which time they cannot transmit. They can then develop asymptomatic infection (entering the *A* class) or symptomatic infection (entering the *I* class). We assume that all asymptomatic individuals will recover (*R*). Those with symptomatic infections can either recover or require hospitalization (entering the *H* class). Some of those hospitalized will die (entering the deceased class *D*), and the rest will recover. Due to the short time scale of our simulations, we do not model births or deaths from non-COVID causes or waning of immunity.

To account for heterogeneity in susceptibility to both infection and severe disease, we further stratify this seven compartment transmission model by both age (*<*20 years, 20-64 years, and ≥65 years) and risk (high vs. low risk). The high risk group was parameterized to capture individuals who are at high risk of infection either because of their occupational exposure level (for example, due to working as a healthcare worker or teacher) or due to underlying health conditions. The size of the high risk group for each age group based on underlying health conditions was estimated based on data from Clark et al [19], and the fraction of individuals with occupational exposure was calculated based on the National Academy of Sciences [20]. The relative risk of infection for those with high risk underlying conditions compared to low risk individuals without these risk factors was estimated based on data from Clark et al. [19]. For simplicity, individuals with high occupational exposure were assumed to have the same relative risk of infection as individuals with underlying health conditions. See SI for relative parameter values.

### Baseline immunity by age group

We assume that the level of baseline immunity is 32%, roughly 4x the number of reported cases in the United States as of February 23, 2021 [1]. As a sensitivity analysis, we also considered simulations with a lower level of starting immunity (16%). For both scenarios, we consider starting baseline immunity to confer protection against infection for the duration of the simulation. While other types and degrees of baseline immunity are possible [21], we focus on this scenario as a reasonable approximation, particularly for the short time scale of our simulations. The age distribution of immunity was calculated based on seroprevalence data from CDC for four states: Georgia, California, Wisconsin, and New York [22]. These four sites were chosen to represent different regions of the country. For the full US model, the age distribution of prior infections (as measured by serological data) was averaged across these sites and assumed to be a proxy for the relative level of immunity for each age group as of early February, 2021. Our calculations accounted for unequal probability of sampling by age based on the 2019 American Community Survey data for each state [23].

### Implementing vaccination

We implement vaccination by adding a daily overall rate of vaccination *λ* and additional compartments for vaccinated individuals, which mirror the compartments in the base model: *S*_*V*_, *E*_*V*_, *A*_*V*_, *I*_*V*_, *R*_*V*_, *H*_*V*_, and *D*_*V*_. We assume that antibody testing will not be used to assess whether an individual has already been exposed, and therefore susceptible and recovered individuals are equally likely to receive the vaccine. For simplicity, we assume that individuals in the other 5 compartments *E, A, I, H*,and *D* will not be vaccinated. Following vaccination, susceptible individuals *S* move into the susceptible vaccinated compartment *S*_*V*_, and all recovered individuals move into the recovered vaccinated compartment *R*_*V*_. A diagram of our model is shown in Figure 1.

**Figure 1:**
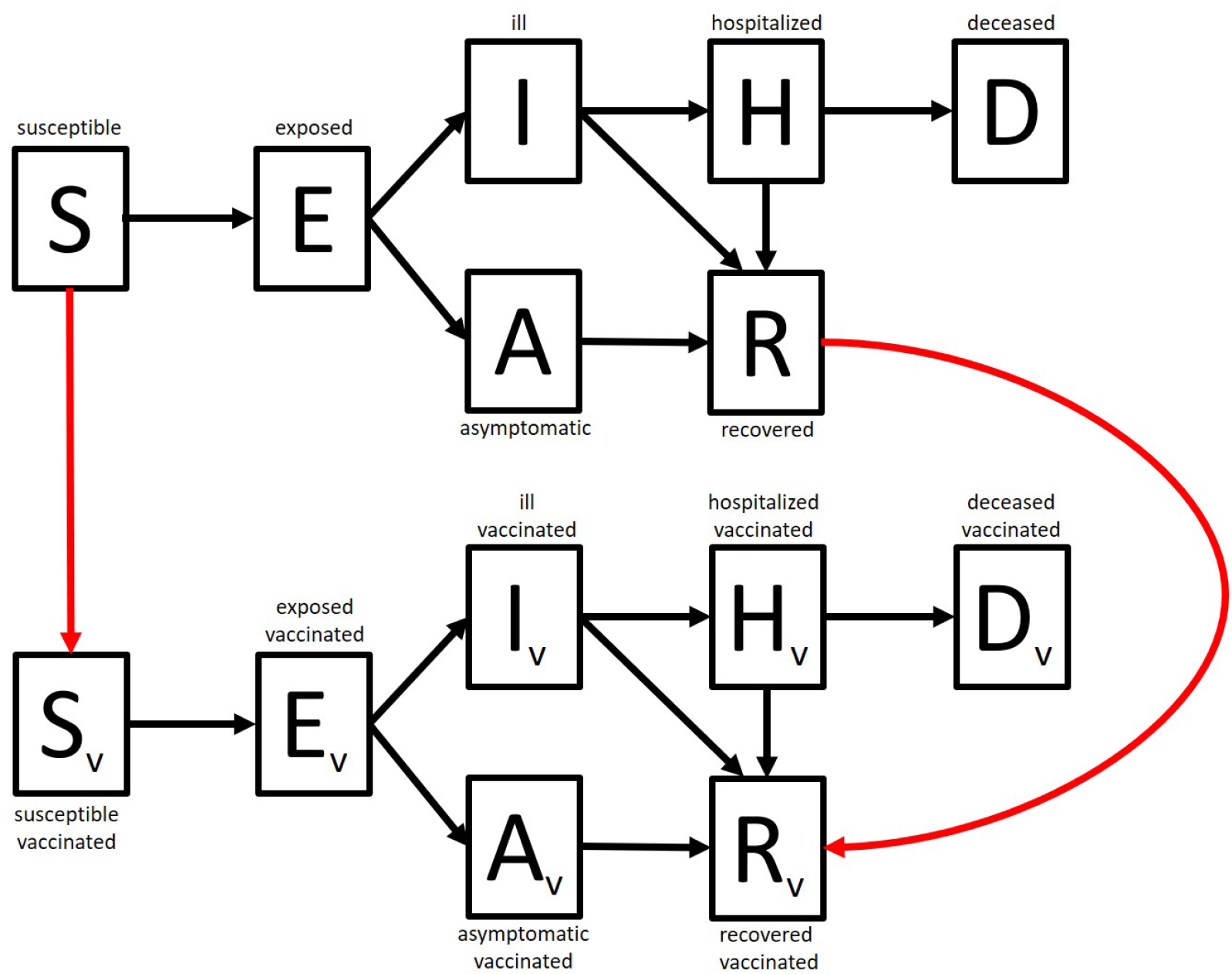
Schematic of the base transmission model with vaccination. Individuals are classified as susceptible, *S*, exposed *E*, infectious and asymptomatic, *A*, infectious and symptomatic, *I*, hospitalized, *H*, recovered, *R*, or deceased, *D*. We assume that recovered individuals are equally as likely as susceptible individuals to receive the vaccine, effectively resulting in wasted doses (red arrows). Vaccinated individuals enter a reduced risk state, in which they are less likely to become infected and may also be less likely to develop symptoms, depending on the modeled mechanism of action.

The rate of vaccination was calculated by assuming that vaccines will preferentially be distributed to high risk groups and to the elderly based on the prioritization scheme proposed by the National Academy of Sciences and the CDC [20, 24]. In our framework, elderly individuals with underlying health conditions have the highest priority followed by low-risk elderly, high risk adults, and low risk adults. Given that children were not included in the initial trials, we do not model vaccination of children. While older teenagers are eligible for vaccination (16-17 year olds could receive the Pfizer vaccine and 18-19 year olds could receive all three vaccines approved for emergency use [6, 5, 4]), we do not model vaccination of this group due to its relatively small size, the initial low rate of vaccine distribution in this age group, and for simplicity. Because identification of high risk groups is likely to be difficult to achieve in real time, we assumed that vaccines would be distributed to all four of these groups (high risk elderly, low risk elderly, high risk adults, low risk adults), with more doses initially being allocated to the higher risk groups. Once vaccine coverage reached 80% in a given group, the remaining doses were redistributed among the remaining vaccine-eligible groups. This 80% coverage threshold is consistent with vaccine acceptance from recent surveys in the United States [25, 26]. Parameter values for the model are shown in Tables S2 and S1.

We model a vaccine that reduces symptomatic infection by 90% (approximately in line with vaccine efficacy of the Pfizer and Moderna mRNA vaccines) in two ways: 1) by reducing infection but not impacting disease progression (*V E*_*susceptibility*_ = 0.9, *V E*_*infectiousness*_ = 0, *V E*_*progression*_ = 0), hereafter referred to as ‘susceptibility-only vaccine’ or by 2) reducing both infection and the probability of symptoms given infection (*V E*_*susceptibility*_ = 0.67, *V E*_*infectiousness*_ = 0, *V E*_*progression*_ = 0.70), hereafter referred to as a ‘susceptibility and severity vaccine.’ These quantities were chosen so that the observed impact on symptomatic disease is the same (i.e., (1 − *V E*_*susceptibility*_) × (1 − *V E*_*progression*_), so (1 − 0.67) × (1 − 0.7) = (1 − 0.9)). We conservatively assume that vaccination does not reduce infectiousness once infected. Given that efficacy against symptomatic COVID-19 is so high, a scenario where vaccination only reduces symptoms but not infection is unlikely, so we did not consider this possibility in our models. Dagan et al recently showed a 90% reduction in COVID-19 infection without recorded symptoms following 2 doses of vaccine, a proxy for asymptomatic infection, consistent with the susceptibility only vaccine [16].

We considered an aggressive vaccination rate of 3 million doses/day as the upper bound of rollout speed (similar to what is achieved for seasonal influenza each season) [27] and a lower bound of 1 million/day, corresponding to the Biden administration’s initial rollout goal [28]. As of March 4, 2021, current data indicate a daily rate of 1.7 million doses/day, intermediate between these two scenarios [1], with plans to scale up vaccine rollout as additional supplies of the newly approved Johnson and Johnson vaccine become available [29].

These two rates were converted to the number of complete two dose series available each month, (dividing by two for a two-dose series). The number of doses was then converted to a time-varying rate of vaccination for each age and risk group. We assume that all individuals age 20 and older will eventually have access to vaccination, such that vaccine coverage will eventually reach 80% in both strategies, but that a one dose strategy can achieve this coverage level more quickly. The efficacy of a one dose vaccine has not been tested at large scale in Phase III clinical trials, so we varied this quantity, assuming the relative efficacy (RE) of 1 dose compared with 2 doses was 100%, 80%, or 60%. We focus on results for an 80% RE vaccine in the main text, as this value is similar to the relative efficacy of the Johnson and Johnson vaccine compared with the two mRNA vaccines 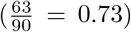and because preliminary trial data have suggested that the 80% threshold is most similar to the level of protection observed between the first and second doses for the Pfizer vaccine in a large field study in Israel 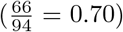 [16]. Additional simulations with lower relative efficacy values are shown in SI.

### Relaxing non-pharmaceutical interventions

Age stratified contact patterns were based on US data from Prem et al. [30]. We assume that at the start of the simulation individuals have 45% of their baseline social contacts, which was calibrated to achieve transmission rates that matched hospitalization data in the United States in early February. This reduction in baseline social contacts is meant to capture not only the reduction in number of interactions, but also their propensity to cause transmission due to mask wearing, surface disinfection, and enhanced hand hygiene. We model the relaxation of NPIs by having all individuals gradually recover more of their social contacts, with the fraction increasing to pre-pandemic levels at a linear rate. We assumed that both vaccinated elderly individuals and all unvaccinated individuals have a delay in relaxation. We vary both the time until relaxation begins for unvaccinated/vaccinated elderly (beginning 0-240 days after vaccination begins) and the time at which pre-pandemic contact rates are restored (for both vaccinated and unvaccinated individuals, ranging from 1 month-1 year after vaccination begins).

For vaccinated individuals, we considered two possibilities. First, we considered what might happen if vaccinated adults began to regain social contacts after receiving their full series instead of waiting a specified number of months. Second, we considered a scenario where vaccinated individuals waited to relax with the general population. In the first scenario, while vaccinated individuals begin to relax NPIs sooner than unvaccinated people, we assume that their social contacts are not fully restored until the general population relaxes due to ongoing restrictions likely to shape social contact patterns. When comparing between relaxation scenarios, we assume that pre-pandemic contact rates are restored at the same time point, such that scenarios in which reopening is delayed have a quicker relaxation process (see Figure 2).

**Figure 2:**
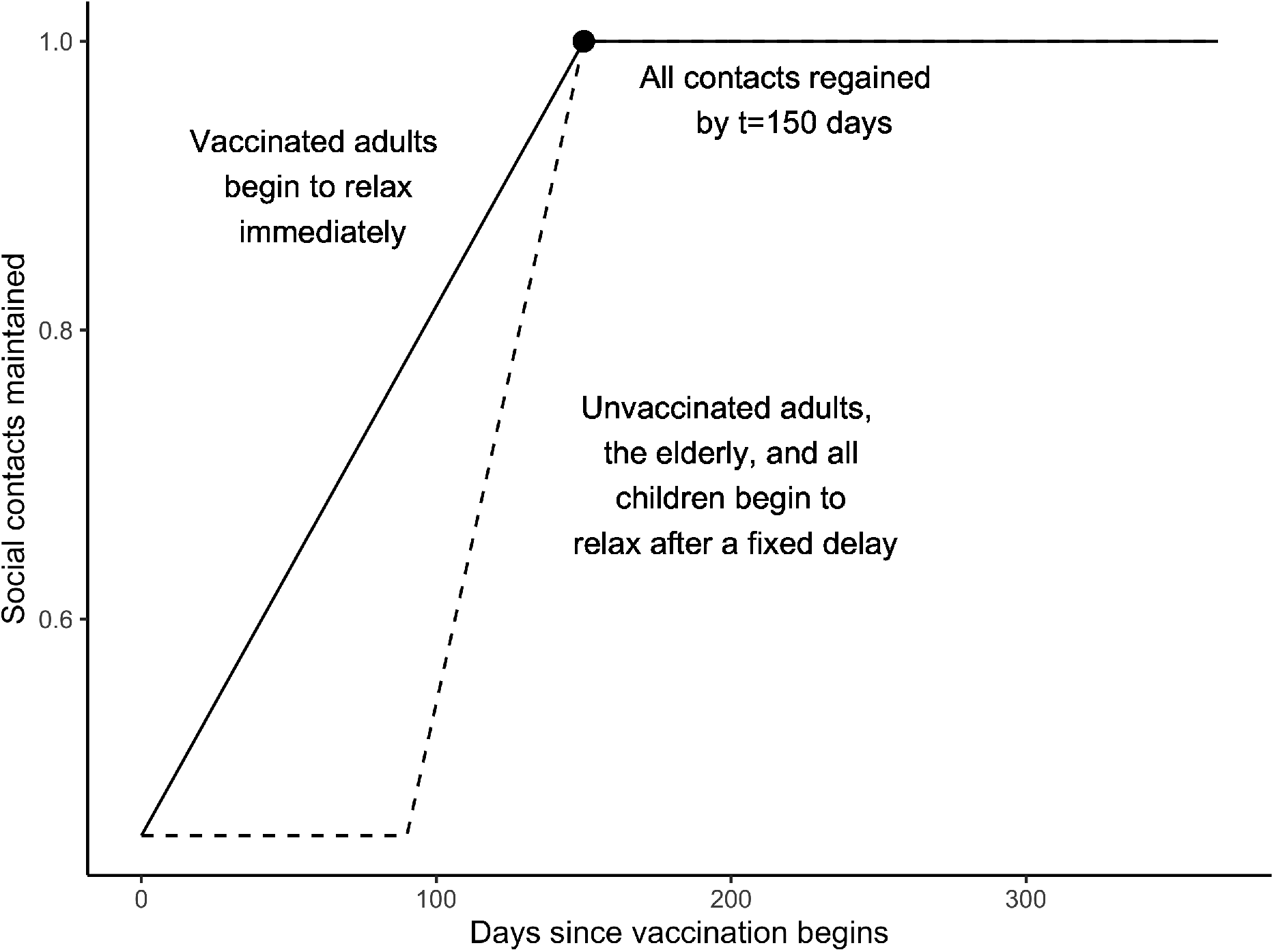
An illustrative example of contact levels over time while relaxing social contacts. Under this scenario, pre-pandemic social contact rates are regained after 150 days for everyone, but the rate of relaxation varies by vaccine status and age. Vaccinated adults (solid line) begin to relax as vaccination begins but unvaccinated individuals and the elderly begin to relax 90 days after vaccination starts (dashed line). Model scenarios considered ranged in both the time at which social contacts were restored to pre-pandemic levels (*t* = 30 to 365) and the initial time at which NPIs started to be relaxed for both vaccinated and unvaccinated individuals (0-240 days).

### Model calibration

We parameterized the model’s reporting rate for symptomatic cases reported and the baseline level of social distancing against US hospitalization data in early February. We found that a reporting rate of 75% combined with a 55% baseline reduction in social contacts closely matched the observed hospitalization data. See Figure S1 for comparisons of incidence between modeled and observed hospitalizations.

## Results

Because relaxing NPIs increases transmission rates (with 100% of social contacts corresponding to an estimated *ℛ*_0_ value of 2.5), the benefits of vaccination in the context of relaxing NPIs depend on both the timescale of relaxation and the speed of vaccine rollout, as both shape the level of population immunity (Figure 3). Relaxing NPIs prematurely, before a substantial fraction of the population has been vaccinated, could lead to dramatic increases in deaths and hospitalizations. For example, beginning to relax NPIs immediately and reaching pre-pandemic contact levels in 30 days, without vaccination, could lead to 1.2 million additional deaths. Even with an effective susceptibility only two dose vaccine, nearly 1 million additional deaths are predicted if NPIs were relaxed over the next month (Figure 3).

**Figure 3:**
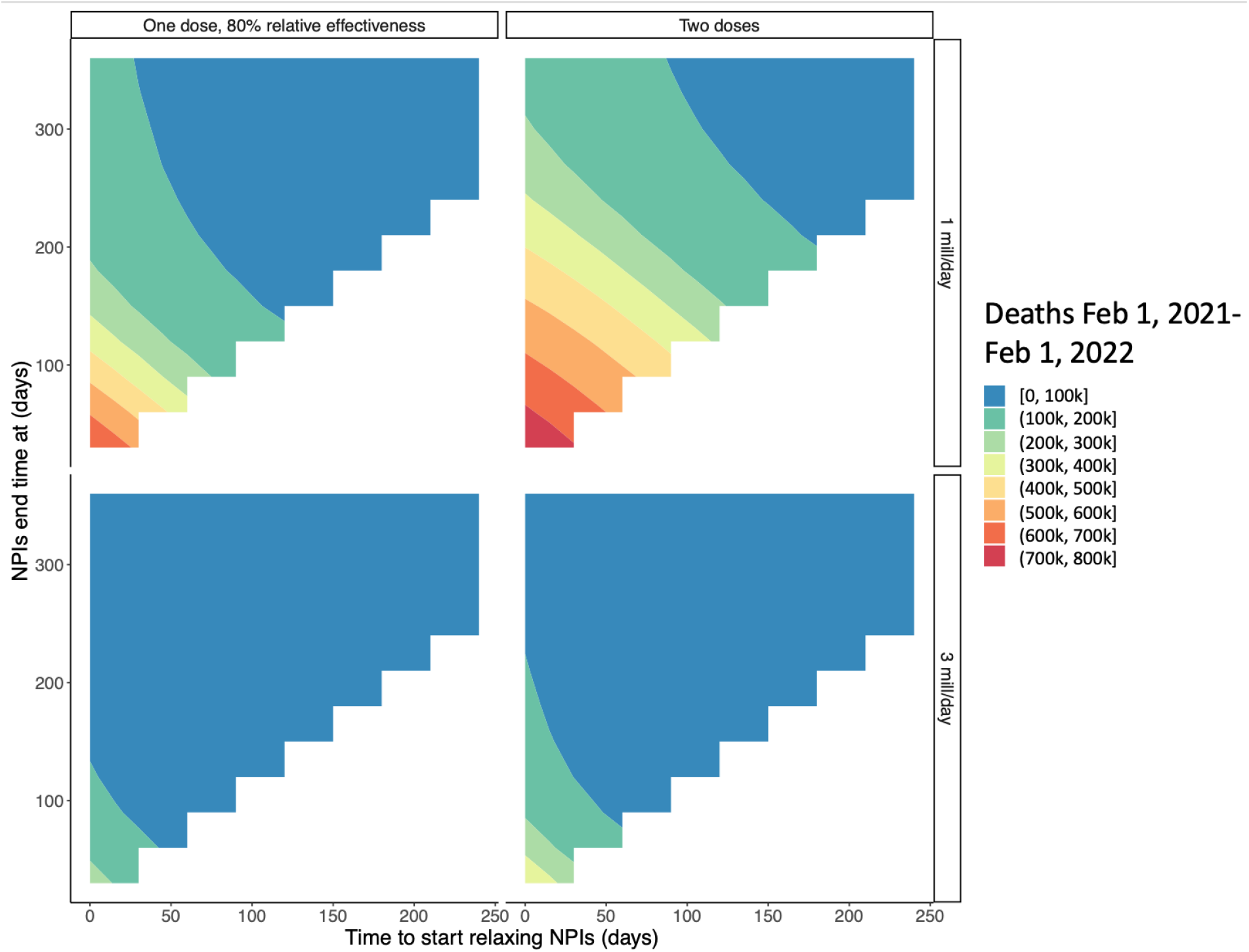
Interaction between time to regain pre-pandemic social interactions and the start time of relaxation by vaccine strategy and rollout speed. The x-axis shows the number of days between February 1, 2021 and the start of further NPI relaxation and the y-axis shows when normal interactions are restored after reopening begins (corresponding to the speed of relaxation). Colors show expected deaths for each reopening strategy. For these simulations, vaccinated individuals are assumed to begin relaxing immediately and a susceptibility only vaccine is modeled. See Figure S4 for expected impacts at lower relative efficacy.

However, delaying reopening for three months while vaccinating rapidly (3 million doses/day) with a susceptibility only vaccine and then rapidly relaxing could reduce deaths to 84,200 and allow social interactions to be fully restored within 3 months. In contrast, due to the slow acquisition of natural immunity with ongoing NPIs, delaying reopening without vaccination results in 1.1 million deaths. Therefore, vaccination can yield a 93% reduction over expected deaths under the same relaxation scale without vaccination. Moreover, in this case, the number of deaths expected are similar to what would be predicted if NPIs were sustained at their current level for the duration of the epidemic (80,000 deaths predicted, see Figure S2), suggesting that early relaxation produces no additional risk. Vaccination also shortens the duration of the epidemic, with incidence falling to near zero within about 3 months with rapid vaccination (Figure 4) compared with about 7 months without vaccination (Figure S3). A one-dose strategy can allow safe relaxation to be achieved more quickly, but could also become a liability if relative efficacy is low (less than 80%) (Figure 3, Figure S4), necessitating a slower relaxation to allow more of the population to be vaccinated. Moreover, speed is less of a benefit if rapid vaccine rollout can be achieved at 3 million doses/day. While waiting for adequate coverage to be achieved before widespread NPI relaxation begins, we found that allowing vaccinated individuals to begin to relax immediately does not substantially increase population transmission (Figure 5). For example, our model predicted using a 2-dose susceptibility only vaccine with a fast rollout, waiting 3 months to relax, and then doing so completely would lead to 84,200 deaths if vaccinated individuals begin to relax sooner, or 83,700 if vaccinated individuals waited to relax with the general population. This slight increase in risk was more pronounced if both a one dose strategy were used and the vaccine was less protective against asymptomatic infection (susceptibility and severity vaccine).

**Figure 4:**
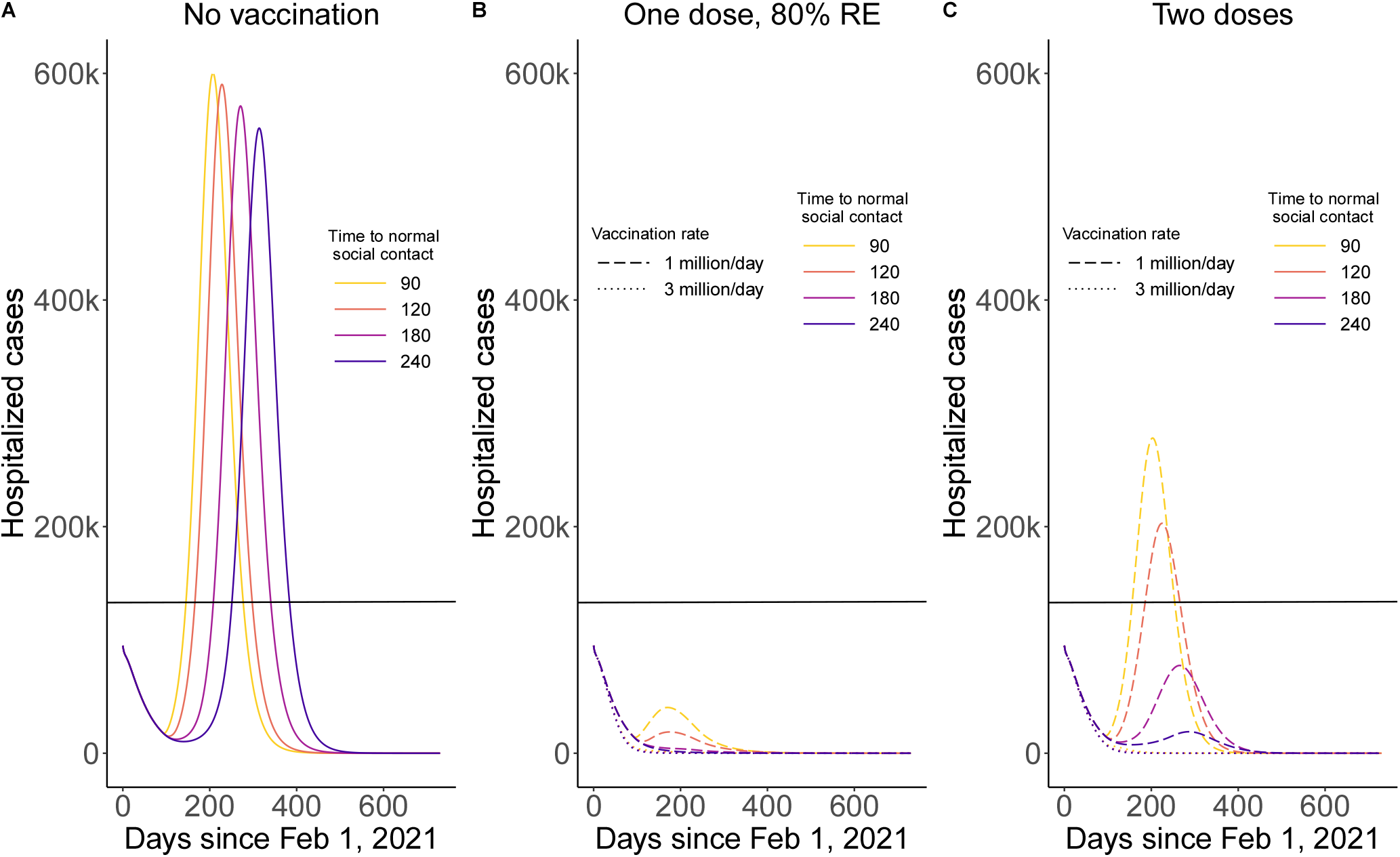
Number of hospitalized cases over time if non-pharmaceutical interventions begin to be relaxed after 90 days based on dosing strategy and speed of reopening. The black line shows the number of individuals hospitalized on January 12, 2021 (131,326), which was the peak of US hospitalizations.

**Figure 5:**
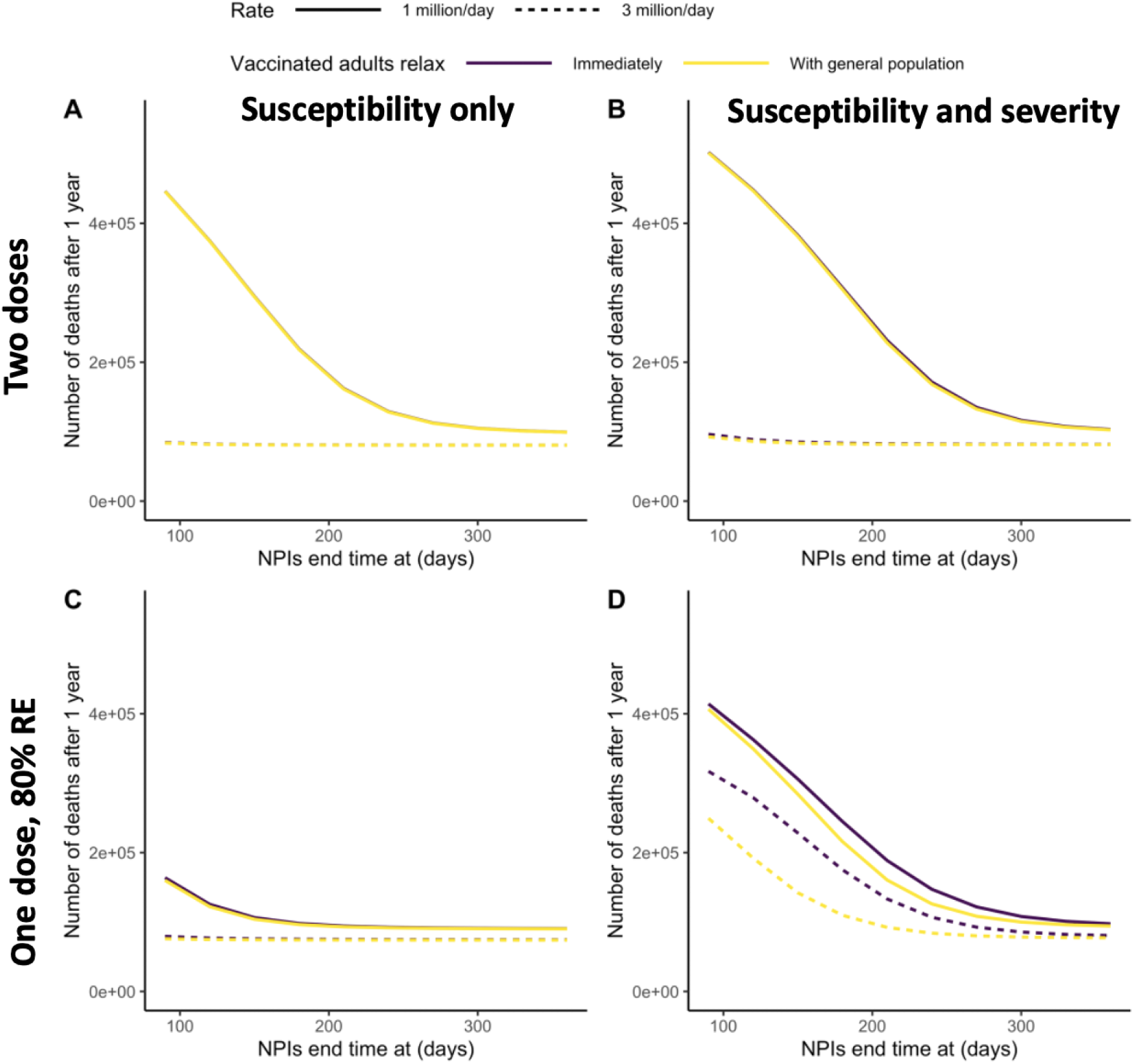
Expected deaths after one year if vaccinated individuals begin to relax immediately (black line) or wait to relax with the general population (yellow line) by the rate of vaccine rollout (solid=1 million doses/day, dashed=3 million doses/day). The top row (A and B) shows impacts for a two dose vaccine and the bottom row (C and D) shows impacts for a one dose vaccine with 80% relative efficacy. Column 1 shows impacts for a susceptibility only vaccine (A and C) and column shows impacts for a susceptibility and severity vaccine (B and D). In panel (A), the black and yellow lines overlap.

The extent of delay and speed of reopening needed to safely relax NPIs for unvaccinated individuals and the elderly depends primarily on the rate of vaccine rollout and the dosing strategy (one or two doses) (Figure 3). If a two dose strategy is used with a rollout rate of 3 million doses/day, a three month delay in reopening provides the most benefit, with a return to normal interactions immediately after 3 months resulting in 84,200 deaths, similar to if NPIs were sustained at their current level of the duration of the epidemic. At this rapid rollout rate, any potential benefit from a one dose strategy is minimal. This three month delay roughly corresponds to the estimated time required to vaccinate 40% of the eligible population, allowing herd immunity to be achieved through a combination of natural and vaccine derived immunity. In contrast, with a slower vaccination rate of 1 million doses/day, the epidemic would not be shortened and NPIs would need to be sustained for the duration of the epidemic, with relaxation not beginning until 7 months after vaccine rollout. A one dose strategy allows high levels of vaccine coverage to be achieved more quickly, but this strategy could be a liability if relative efficacy is low and NPIs are relaxed too quickly (Figure S4). Delaying relaxation also allows NPIs to be relaxed more quickly without increasing population risk. For example, if relaxation began immediately, it would need to be prolonged over the course of the next year to maintain low levels of incidence, whereas NPIs could be relaxed very rapidly with a modest delay of at least 3 months if the rate of vaccine rollout is fast.

Delaying reopening and prioritizing the speed of vaccine rollout can also help reduce the burden on the US healthcare system, preventing or reducing a second wave of hospitalizations. If relaxation begins immediately or the vaccine primarily protects against severe disease rather than infection, a new wave of hospitalizations is expected unless vaccine rollout can be achieved at 3 million doses/day and a two dose strategy is used (Figure S5, Figure S6). Moreover, this wave of hospitalizations would be expected to exceed the levels of burden seen in early January 2021 at the slower rollout speed unless the rate of relaxation is slow, even with a two dose strategy. However, if relaxation is delayed for unvaccinated individuals for 3 months, a second wave of hospitalizations can be prevented if vaccination can be rolled out quickly and vaccine efficacy is high or if vaccine rollout is slower and relaxation occurs gradually over a 3-5 month period (Figure 4). Delays greater than 3 months can allow NPIs to be relaxed more quickly once reopening occurs without risking a second wave.

If baseline immunity is lower than we have modeled, a slightly longer delay in relaxation would be needed (Figure S7). For a two dose vaccine with fast rollout, a 3 month delay brought expected additional deaths to under 178,000, but waiting an additional 2 months (reopening after 150 days) could save an additional 62,000 lives, dropping expected deaths to 116,000. Delays less than 3 months led to dramatic increases in expected deaths, with a complete reopening after 60 days leading to 314,000 deaths. If vaccine rollout was only 1 million doses/day, a delay of at least 7 months was needed to minimize deaths.

## Discussion

Widespread vaccination has the potential to greatly reduce the adverse health consequences of the COVID-19 pandemic and allow a quicker return to normal social interactions with relative safety. If NPIs were relaxed quickly and no vaccination were used, 1.2 million more deaths over the next year would be expected, with a high level of ongoing transmission occurring throughout the next year. In contrast, vaccination with a two dose series coupled with a delayed relaxation of NPIs could reduce deaths by up to 93% (with expected additional deaths between February 1, 2021 and February 1, 2022 falling to 84,200), and the epidemic largely being over within 3 months. This shortening of the epidemic depends on a rapid rate of vaccination, with a substantially prolonged epidemic being expected if only 1 million doses/day can be achieved. For example, with a 90 day delay in reopening and a slow reopening speed, transmission would be ongoing over the next year, even if a two dose strategy is used.

Compared with beginning to relax NPIs immediately, a delay of at least 3 months could greatly enhance the impact of vaccination, with reopening speed playing a smaller role. Society at large may consider a more rapid reopening after a modest delay optimal, given the social, economic, and public health implications of a sustained shutdown. Delaying reopening allows time to improve treatment, which could reduce mortality. Preliminary evidence suggests that case fatality rates have already begun to decline [31], and preserving health care capacity through a controlled reopening plan can help this pattern continue.

Moreover, even while delaying widespread reopening, our model suggests that allowing vaccinated individuals to begin relaxing social distancing as soon as they complete their vaccine series poses limited population risk, and could allow a substantial fraction of the population to return to normal activities more quickly, alleviating some of the social and economic costs of delaying reopening further. In practice, given that front-line workers have been prioritized for vaccination, allowing these individuals to relax first might lead to further reductions in risk than we have modeled if these individuals ultimately constitute more of social interactions, serving as immune shields [32, 33].

Additional interventions might enable the US epidemic to end sooner, ultimately enabling a quicker return to normal activity levels. We have not modeled vaccination of children because they have so far not been included in clinical trials [34]. However, if pediatric vaccination is shown to be safe and effective, child vaccination could also allow herd immunity to be reached more quickly and for it to be sustained in the long term. Other NPIs could also enhance the potential benefits of vaccination. For example, increased nationwide mask usage might also be contributing to the observed decline in cases [35].

While the indirect effects of COVID-19 vaccines are uncertain, we find that the expected impacts of vaccination are similar when impacts on transmission were at least moderate (70% reduction in susceptibility). Given the high level of impact against symptomatic COVID-19, we expect that there will be a degree of protection against infection. If protection against infection is far lower, the potential benefits of vaccination will be reduced. Post-licensure studies could help provide clarity on the mechanism of action for approved vaccines, including whether or not vaccination reduces infectiousness or susceptibility to infection, either of which would provide indirect benefits to unvaccinated individuals [13, 14]. Already, preliminary evidence from the Pfizer vaccine suggests that effectiveness against asymptomatic SARS-CoV-2 is high and may be similar to impacts observed for symptomatic infection [16].

Two crucial factors might have led us to overestimate the total impact of vaccination. First, if new strains of COVID-19 emerge against which available vaccines are not protective, vaccine impact could be reduced. Additionally, even if vaccination is effective, if these strains are more transmissible, ongoing NPIs might be needed for longer to prevent surges in cases and vaccine coverage might need to be higher before restrictions can be safely relaxed. Some have cautioned that pursuing a one-dose strategy might increase the risk of evolution of vaccine-resistant strains, but this concern remains speculative [36]. As of early March 2021, at least three new variants of concern have emerged, including the UK strain, the South African strain, and the Brazilian strain. At present, all three vaccines approved for emergency use appear to be protective against these strains, but possibly to a lesser extent than the initial variants circulating at the time of vaccine development [37, 38]. Follow-up studies are planned to determine vaccine effectiveness against these and potentially future variants of concern. However, immune escape is still a distinct possibility.

Second, due to a lack of data and the short time scale of our simulations, we did not account for waning immunity in our models either for natural infection or for vaccination. Depending on the duration and degree of immunity, additional waves of transmission are possible, particularly if many individuals choose not to become vaccinated [39]. If immunity is relatively short, high vaccination rates combined with follow up booster doses for vaccinated individuals might be necessary to prevent future transmission waves [39].

In conclusion, we have found that widespread vaccination has the potential to reduce deaths from COVID-19, lessen health system strain, and shorten the length of the COVID-19 pandemic, even as non-pharmaceutical interventions continue to be relaxed. Based on currently available data, we find that using the full recommended series for approved vaccines is likely to make a substantial impact. A one dose series could outperform a two dose series, but if vaccine rollout can be achieved at 3 million doses/day, the additional benefit of a one dose strategy is minimal and the potential risk is high, particularly if VE is low. Using this strategy, a three month delay in further reopening followed by gradual relaxation provides the best opportunity to minimize deaths, with normal interactions being restored within 4 months. Our findings support quick return to normal social interactions for vaccinated people. Additional data on the indirect effects of vaccination and the extent and duration of naturally-acquired and vaccine-derived immunity are urgently needed to help guide policy at this critical stage of the US epidemic. Monitoring circulating strains of COVID-19 as well as vaccine efficacy against them is also critical to ensure ongoing efficacy and determine if vaccine reformulation will be necessary.

## 1 Supplementary information

### 1.1 Model equations

In the equations below, the subscript *i* denotes age group, the subscript *j* denotes risk group (high or low) and the subscripts *vax* and *nv* indicate equations for vaccinated and unvaccinated individuals, respectively.

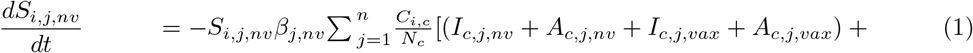

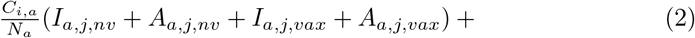

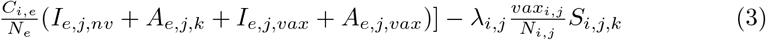

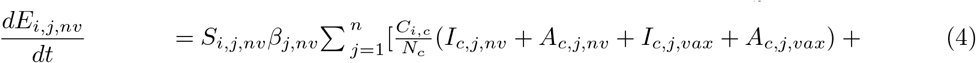

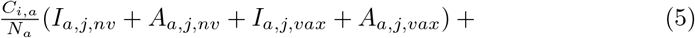

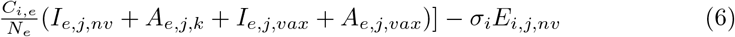

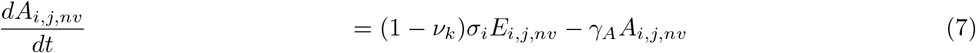

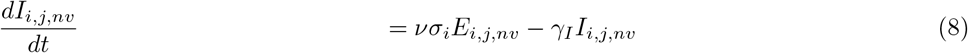

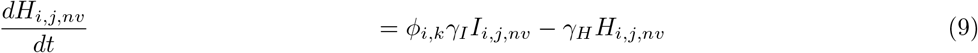

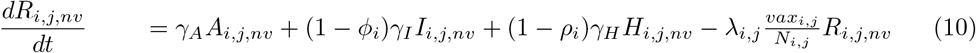

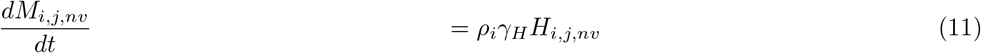

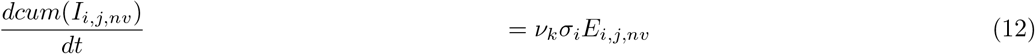

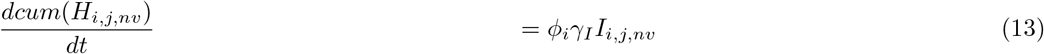

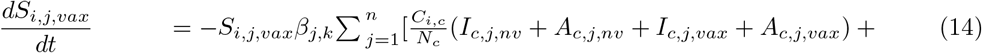

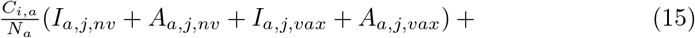

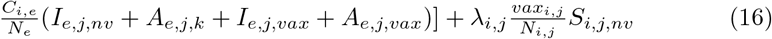

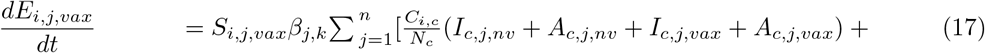

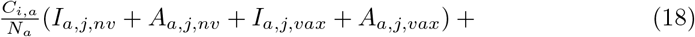

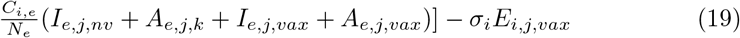

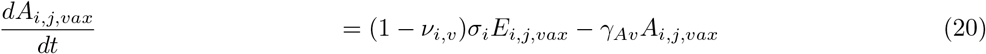

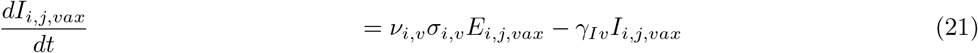

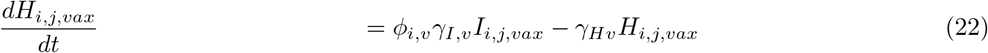

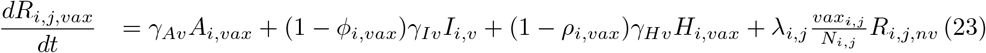

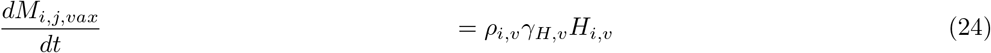

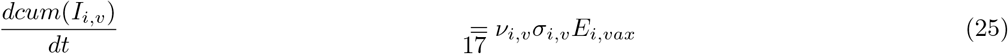

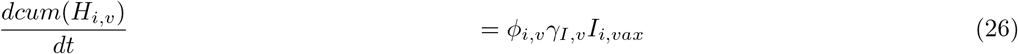

### 1.2 Model parameters

#### 1.2.1 Overall model parameters

##### Parameter values

**Table S1:**
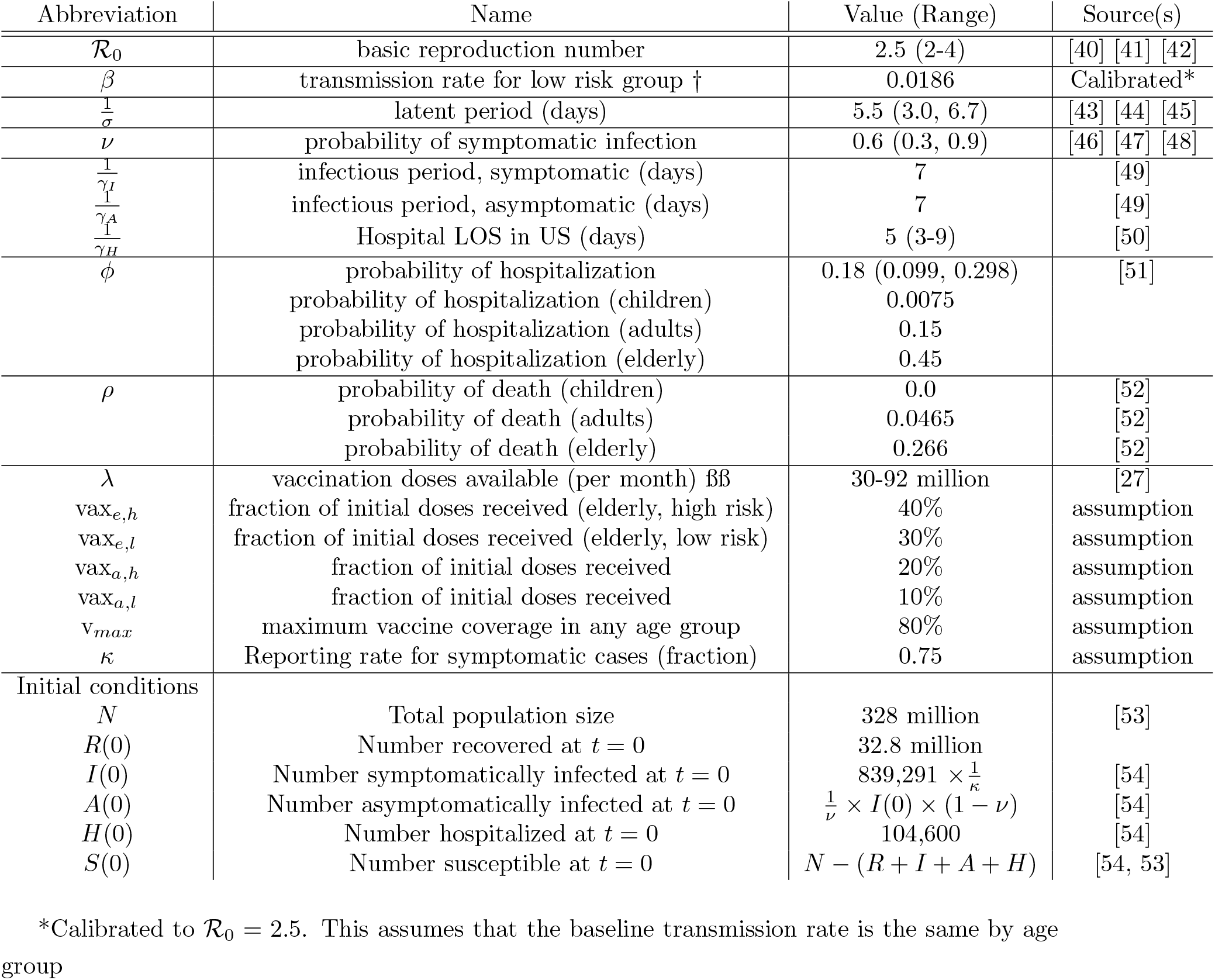
Parameter values for the model are informed by the current best estimates in the SARS-CoV-2 literature.

| Abbreviation | Name | Value (Range) | Source(s) |
| --- | --- | --- | --- |
| $\mathcal{R}_0$ | basic reproduction number | 2.5 (2-4) | [40] [41] [42] |
| $\beta$ | transmission rate for low risk group † | 0.0186 | Calibrated* |
| $\frac{1}{\sigma}$ | latent period (days) | 5.5 (3.0, 6.7) | [43] [44] [45] |
| $\nu$ | probability of symptomatic infection | 0.6 (0.3, 0.9) | [46] [47] [48] |
| $\frac{1}{\gamma_I}$ | infectious period, symptomatic (days) | 7 | [49] |
| $\frac{1}{\gamma_A}$ | infectious period, asymptomatic (days) | 7 | [49] |
| $\frac{1}{\gamma_H}$ | Hospital LOS in US (days) | 5 (3-9) | [50] |
| $\phi$ | probability of hospitalization | 0.18 (0.099, 0.298) | [51] |
|  | probability of hospitalization (children) | 0.0075 |  |
|  | probability of hospitalization (adults) | 0.15 |  |
|  | probability of hospitalization (elderly) | 0.45 |  |
| $\rho$ | probability of death (children) | 0.0 | [52] |
|  | probability of death (adults) | 0.0465 | [52] |
|  | probability of death (elderly) | 0.266 | [52] |
| $\lambda$ | vaccination doses available (per month) ‡ | 30-92 million | [27] |
| $\text{vax}_{e,h}$ | fraction of initial doses received (elderly, high risk) | 40% | assumption |
| $\text{vax}_{e,l}$ | fraction of initial doses received (elderly, low risk) | 30% | assumption |
| $\text{vax}_{a,h}$ | fraction of initial doses received | 20% | assumption |
| $\text{vax}_{a,l}$ | fraction of initial doses received | 10% | assumption |
| $\text{v}_{max}$ | maximum vaccine coverage in any age group | 80% | assumption |
| $\kappa$ | Reporting rate for symptomatic cases (fraction) | 0.75 | assumption |
| Initial conditions |  |  |  |
| $N$ | Total population size | 328 million | [53] |
| $R(0)$ | Number recovered at $t = 0$ | 32.8 million | |
| $I(0)$ | Number symptomatically infected at $t = 0$ | $839,291 \times \frac{1}{\kappa}$ | [54] |
| $A(0)$ | Number asymptotically infected at $t = 0$ | $\frac{1}{\nu} \times I(0) \times (1 - \nu)$ | [54] |
| $H(0)$ | Number hospitalized at $t = 0$ | 104,600 | [54] |
| $S(0)$ | Number susceptible at $t = 0$ | $N - (R + I + A + H)$ | [54, 53] |

\*Calibrated to $\mathcal{R}_0 = 2.5$ . This assumes that the baseline transmission rate is the same by age group

#### 1.2.2 Model stratification

The relative size of each age group was estimated using the US Census data. Within each age group, the fraction of the population considered occupationally exposed was estimated based on the NAP report for prioritization of frontline workers. Both the fraction of the population with high levels of susceptibility and the relative risk of infection for high susceptible individuals was taken from Clark et al. To produce an upper bound on the size of the high risk group, the size of these two groups were summed (used in model runs in the main text). The lower bound can be estimated by assuming that all high susceptibility individuals are also occupationally exposed. There is no difference between these two assumptions for the elderly or for children (because neither age group is occupationally exposed).

**Table S2:** Parameter values by group

| Parameter | Age group |  |  |
| --- | --- | --- | --- |
| | Children (<20 years) | Adults (20-64 years) | Elderly ($\geq 65$ years) |
| Fraction of overall population in age group<sup>1</sup> | 18.7% | 68.7% | 12.6% |
| Fraction high susceptibility<sup>2</sup> | 2.5% | 13.5% | 75% |
| Relative risk of severe disease high susceptibility vs. low (reference group)<sup>2</sup> | 2.0 | 2.55 | 2.55 |
| Fraction occupationally exposed<sup>3</sup> | 0 | 23% | 0 |
| Fraction in high risk group | 2.5% | 23.0%-36.5% | 75% |

1. Based on US Census population in 2019 [53]
2. Parameters taken from Clark et al, 2020 for the US using the spreadsheet tool [19]
3. Based on NAP report for prioritization for front-line workers[20]

### 1.3 Model calibration

Overall, the modeled hospitalizations were consistent with observed data in late February. For model calibration, we assumed the higher bound of baseline immunity (32%) and one million doses of vaccine per day.

### 1.4 Relative one-dose efficacy

Where one dose efficacy was less than 100%, the corresponding efficacy values were calculated separately for a susceptibility only and a susceptibility and severity vaccine. For the susceptibility and severity vaccine, *V E*_*susceptibility*_ and *V E*_*progression*_ were calculated as follows, by relative efficacy (*RE*) values:

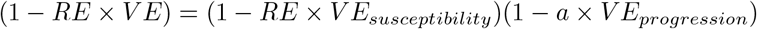

**Figure S1:**
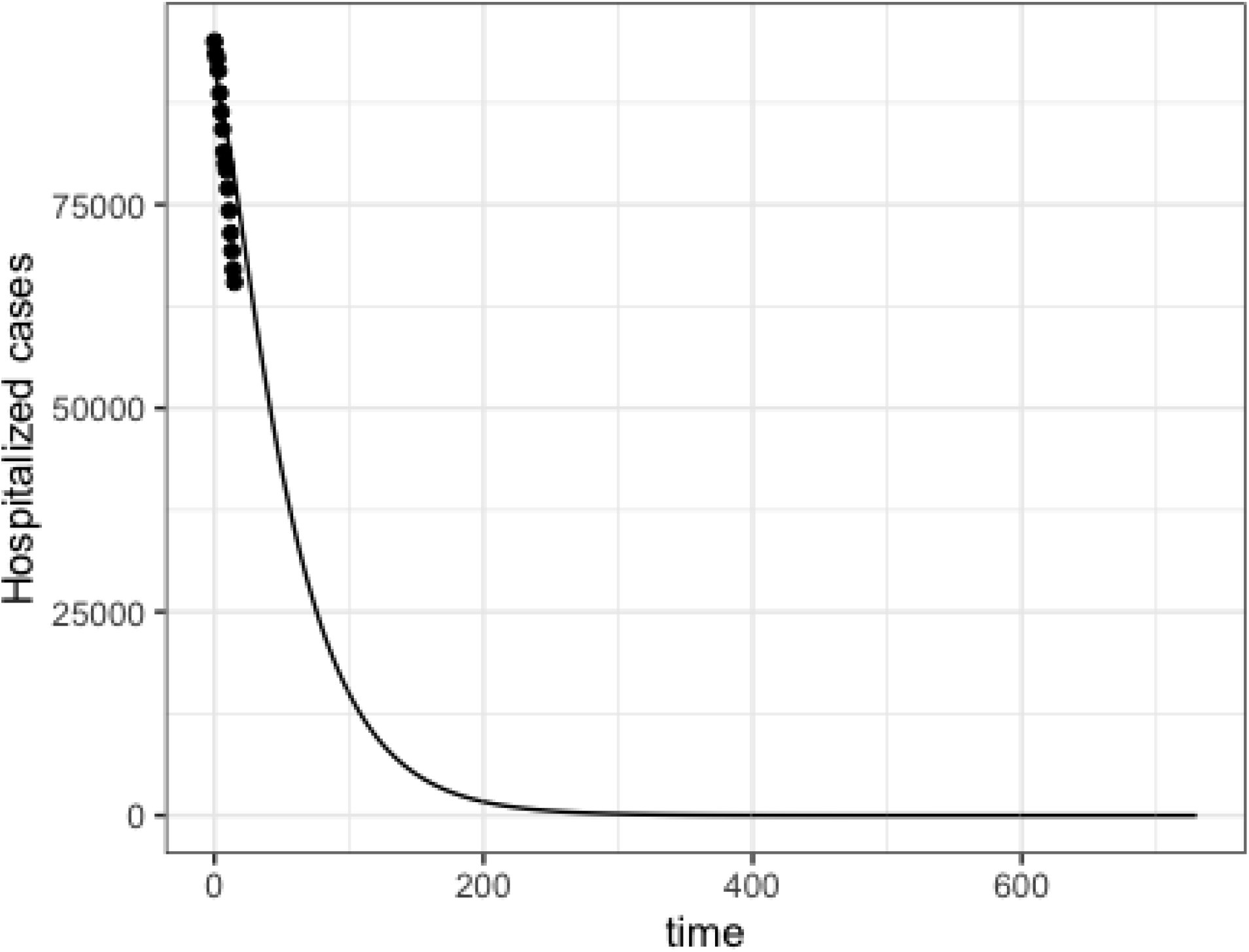
Consistency between modeled hospitalizations (black line) and observed data (points) from the Covid Tracking Project [1]

**Figure S2:**
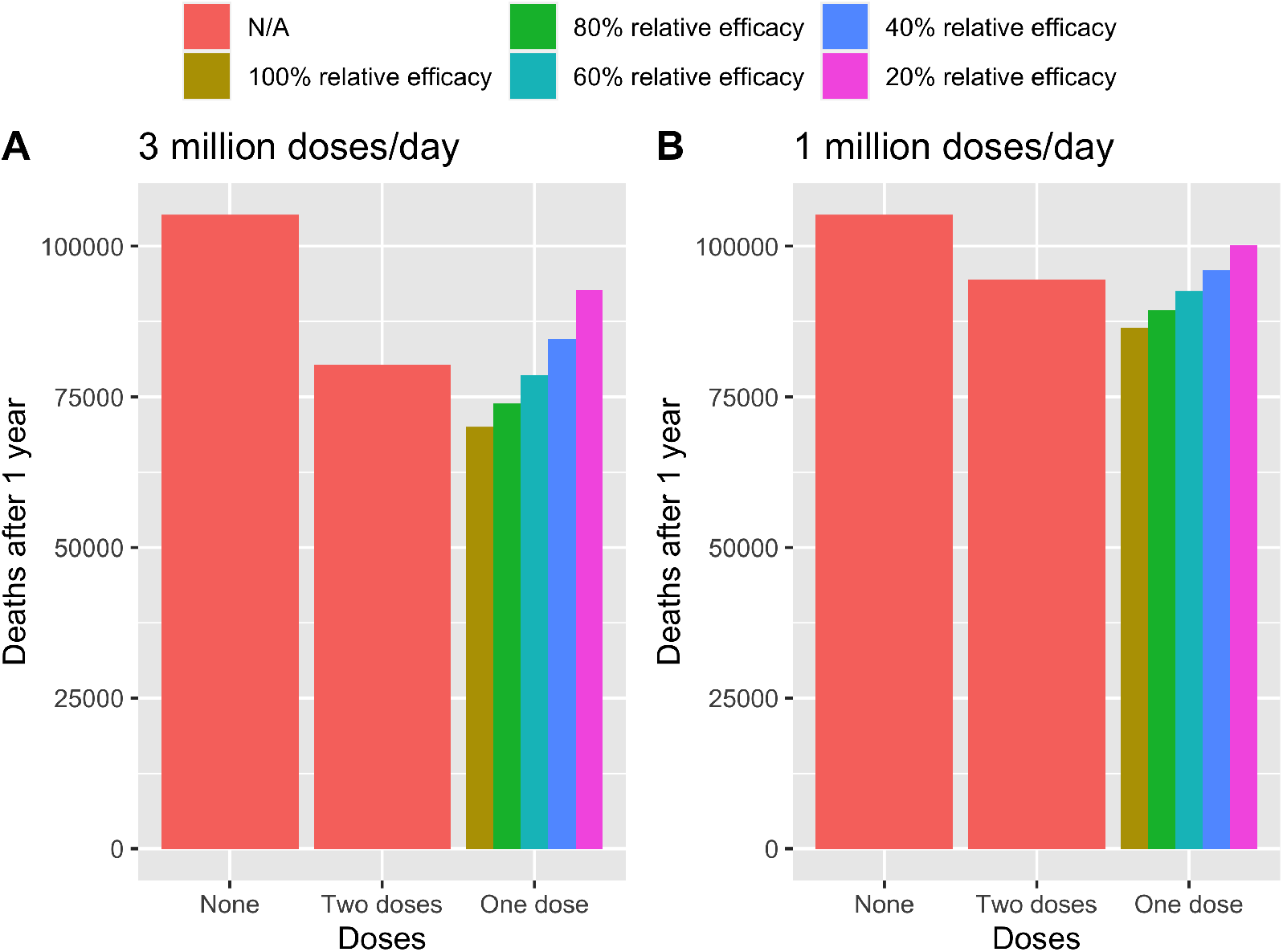
Deaths between February 1, 2021 and February 1, 2022 for different vaccination strategies and relative performance with no relaxation of NPIs. This figure shows performance for a susceptibility only vaccine, but results for a susceptibility and severity vaccine were similar.

We then solved for the value of *a* that satisfied the equation to get the appropriate *V E* values. In the main text, we focus on a one dose vaccine with an 80% relative efficacy, as this is similar to initial data for Pfizer [16].

### 1.5 Hospitalization with constant NPIs

### 1.6 Sensitivity analyses

In sensitivity analysis, we assessed the sensitivity of model conclusions to: 1) the level of baseline immunity, 2) lower RE values, 3) the extent of delay in reopening, and 4) vaccine mechanism of action.

**Figure S3:**
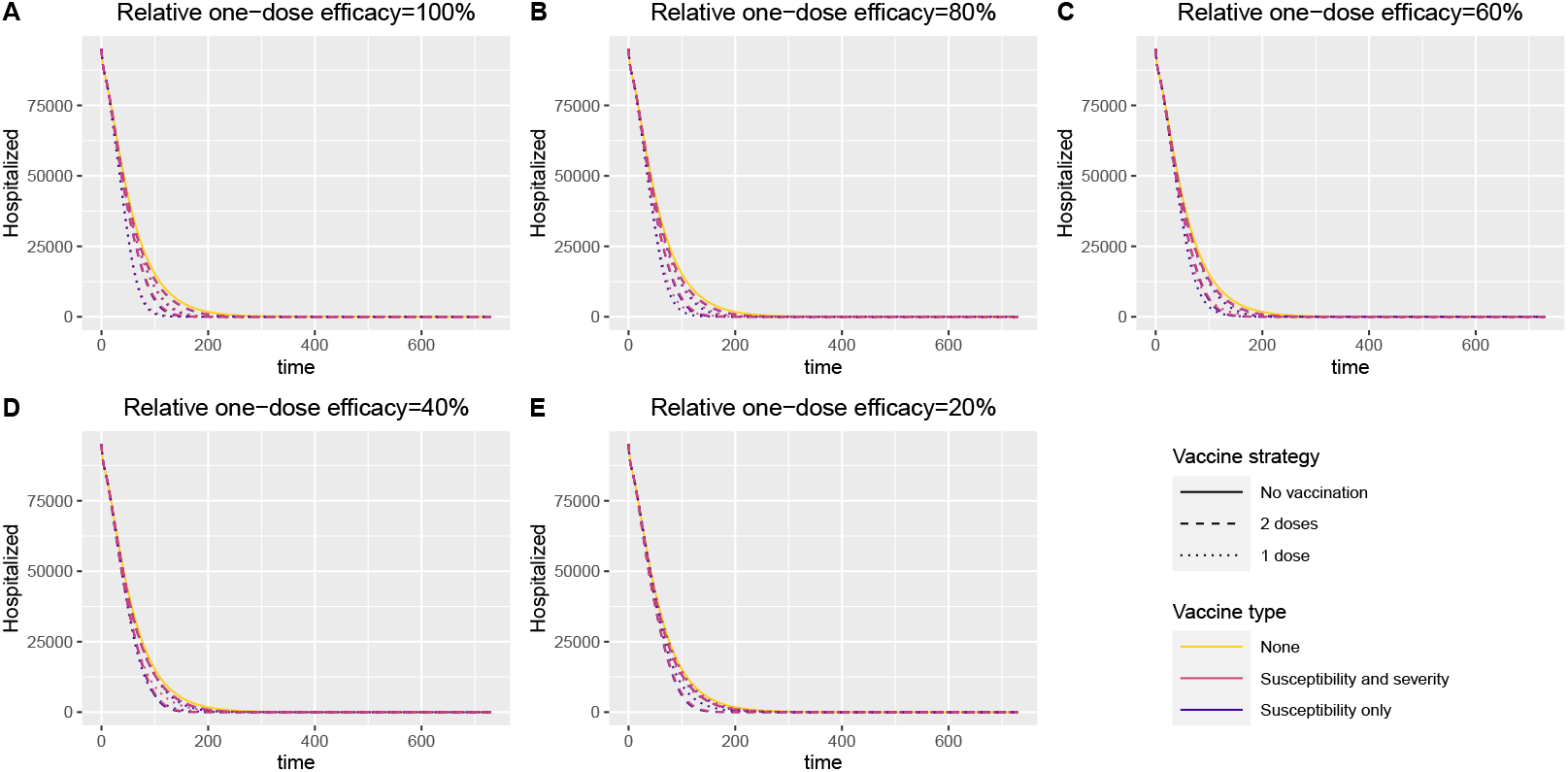
Predicted hospitalizations if NPIs are sustained at their current level for the duration of the simulation (2 years)

#### 1.6.1 Varying RE value

To illustrate the impact of relaxation under different RE scenarios, we re-ran the heat map in the main text showing results for 100% and 60% relative efficacy. In general, results were similar, but a 60% relative efficacy vaccine requires a longer delay before further relaxation begins, with a delay of 4-7 months being needed, depending on the rate of vaccination.

#### 1.6.2 Hospitalizations if no delay in reopening

Without delaying reopening, the model predicts that the US healthcare system would quickly surpass levels seen in early January 2021 at the peak health system load unless vaccination can be rolled out at 3 million doses/day or relaxation occurred gradually over a 6-12 month period.

**Figure S4:**
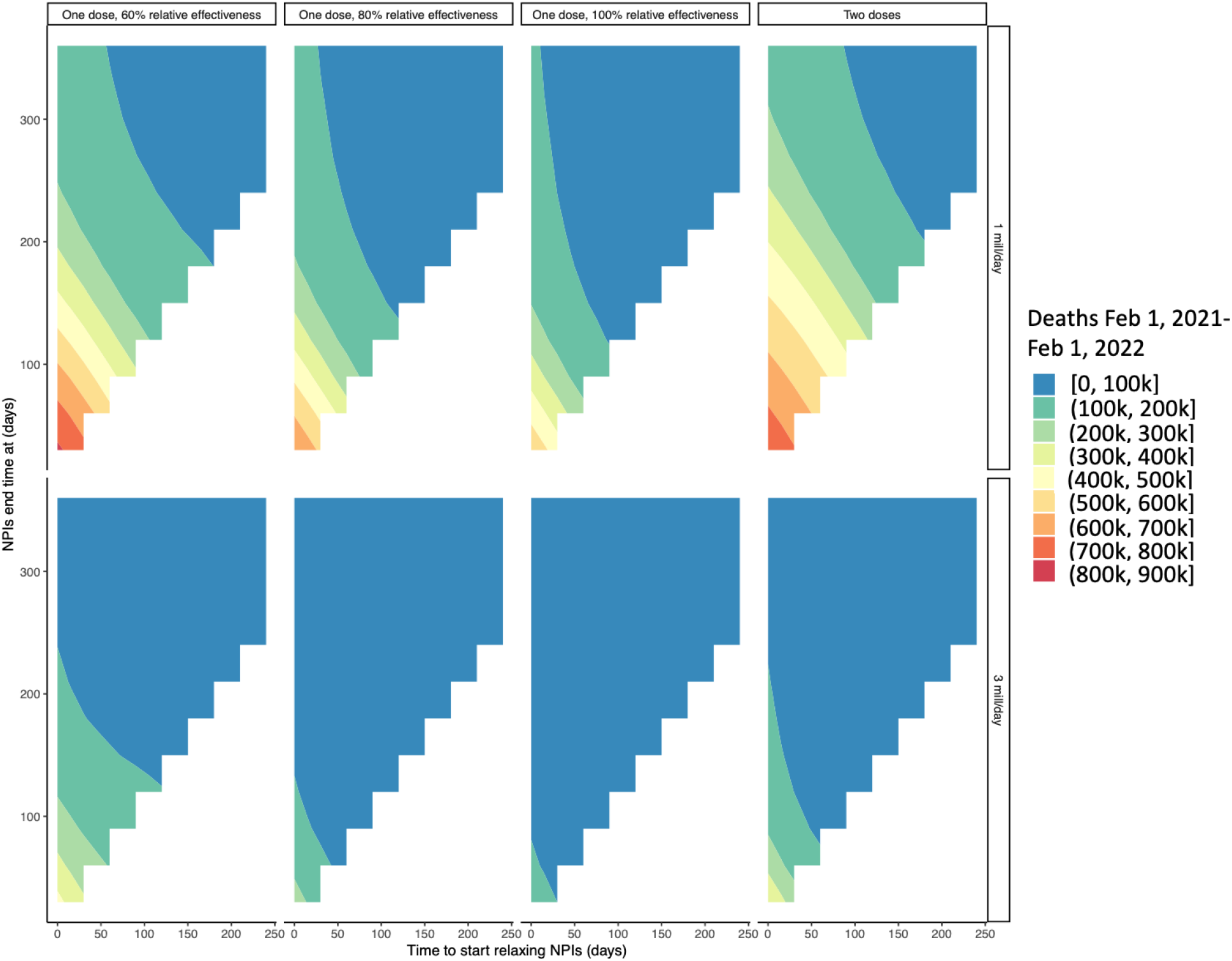
Relaxation tradeoff for varying levels of relative efficacy and rollout speeds for a susceptibility only vaccine.

**Figure S5:**
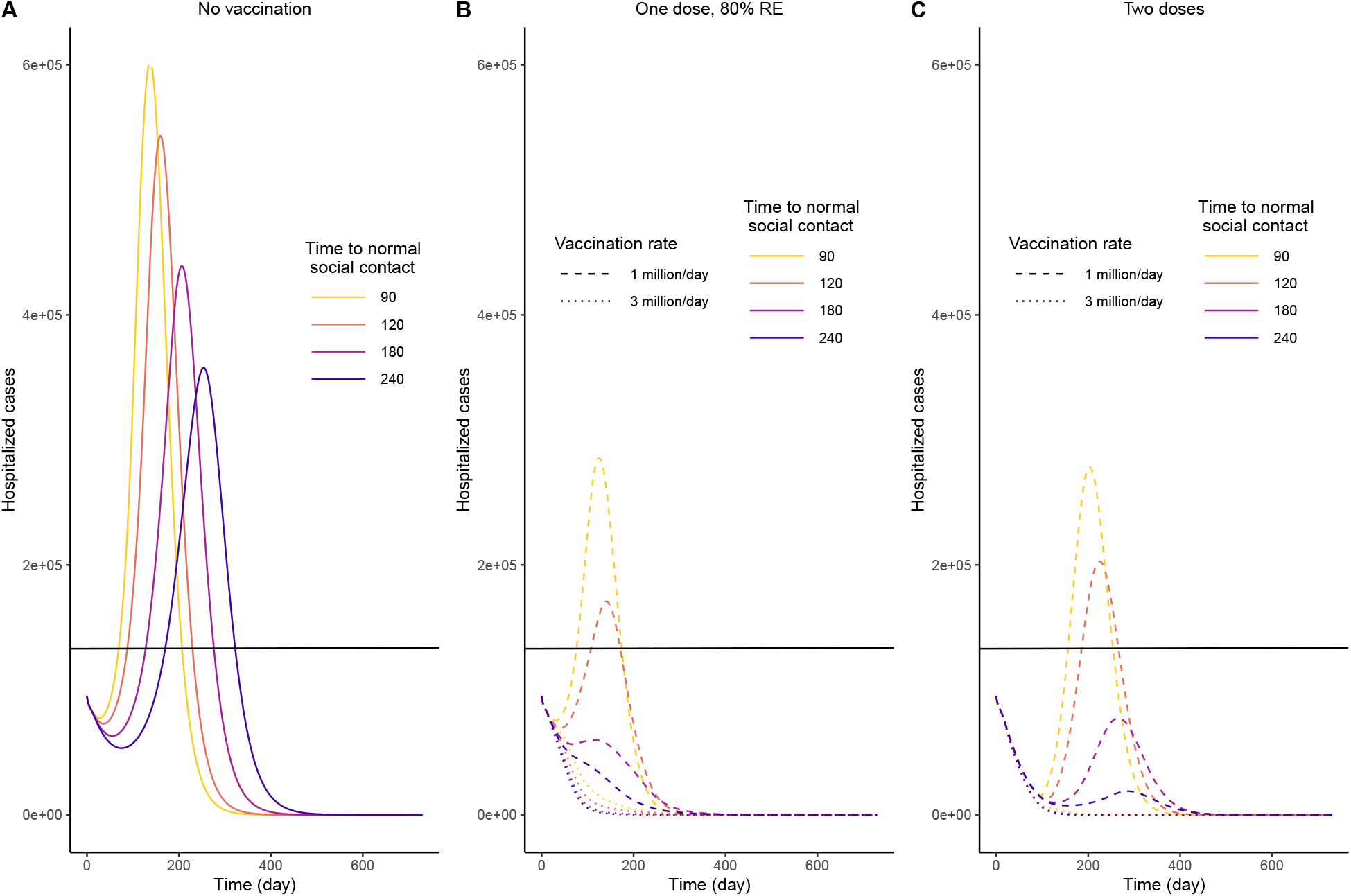
Number of hospitalized cases over time if non-pharmaceutical interventions begin to be relaxed for everyone of February 1, 2021 based on dosing strategy and speed of reopening for a susceptibility only vaccine. The black line shows the number of individuals hospitalized on January 12, 2021 (131,326), which was the peak of US hospitalizations.

**Figure S6:**
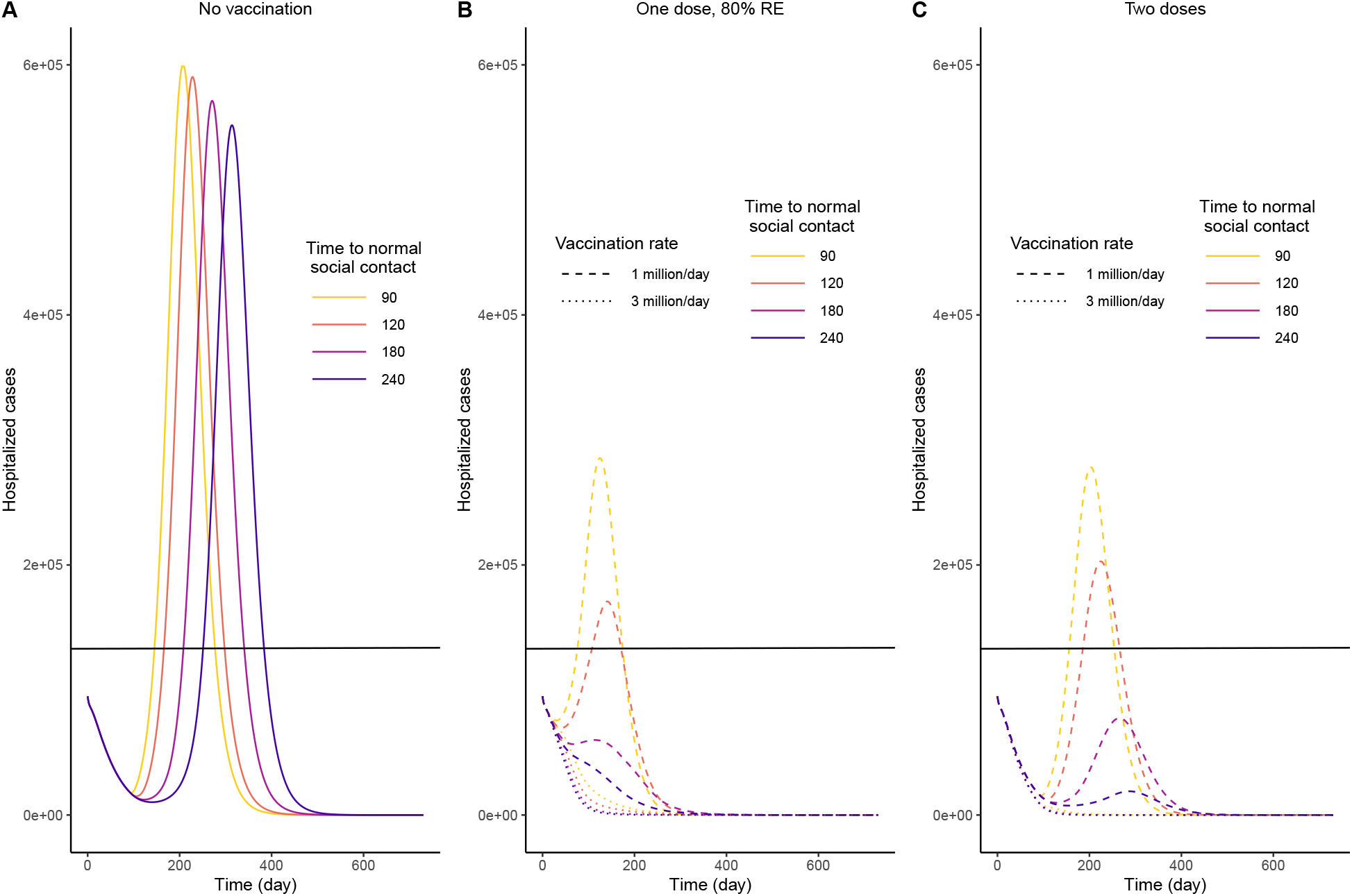
Number of hospitalized cases over time if non-pharmaceutical interventions begin to be relaxed for unvaccinated individuals after 90 days based on dosing strategy and speed of reopening for a susceptibility and severity vaccine. Vaccinated individuals relax as soon as they complete their vaccine series. The black line shows the number of individuals hospitalized on January 12, 2021 (131,326), which was the peak of US hospitalizations.

**Figure S7:**
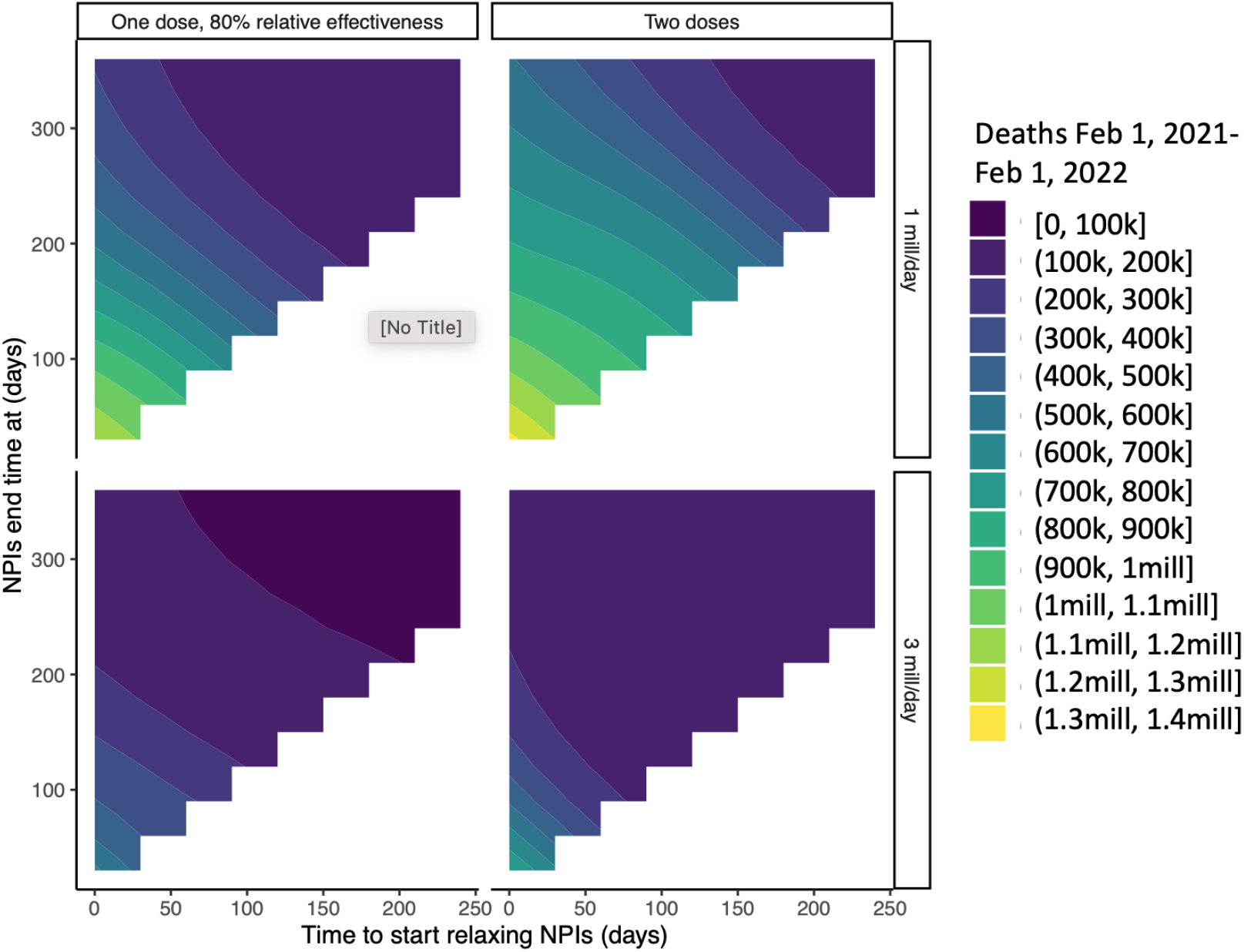
Relaxation tradeoff for deaths for varying relaxation speeds for a susceptibility only vaccine assuming baseline immunity is 16%.

#### 1.6.3 Susceptibility and severity vaccine

Similar to reopening without a delay, the model predicts that if protection against asymptomatic infection is less pronounced, the US healthcare system would quickly surpass levels seen in early January at the peak health system load unless vaccination can be rolled out at 3 million doses/day or relaxation occurred gradually over a 6-12 month period.

#### 1.6.4 Lower baseline immunity (16%)

With a lower level of baseline immunity, the delay needed to minimize deaths is slightly longer, and expected deaths are higher.

## Data Availability

All data used in the manuscript are publicly available. Time series data of COVID hospitalizations used to calibrate the model are available from the COVID tracking project at https://covidtracking.com. Serological data are publicly available on CDC’s website at https://covid.cdc.gov/covid-data-tracker/#national-lab. Census data used to initialize the model are also publicly available at: https://www.census.gov/data/tables/2018/demo/age-and-sex/2018-age-sex-composition.html

